# In the Midst of a Pandemic, Introverts May Have a Mortality Advantage

**DOI:** 10.1101/2022.05.24.22275508

**Authors:** Dana A. Glei, Maxine Weinstein

## Abstract

Extroverts may enjoy lower mortality than introverts under normal circumstances, but the relationship may be different during an airborne pandemic when social contact can be deadly. We used data for midlife Americans surveyed in 1995-96 with mortality follow-up through December 31, 2020 to investigate whether the association between extroversion and mortality changed during the COVID-19 pandemic. We hypothesized that excess mortality during the pandemic will be greater for extroverts than for introverts. Results were based on a Cox model estimating age-specific mortality controlling for sex, race/ethnicity, the period trend in mortality, and an additional indicator for the pandemic period (Mar-Dec 2020). We interacted extroversion with the pandemic indicator to test whether the relationship differed between prepandemic and pandemic periods. Prior to the pandemic, extroversion was associated with somewhat lower mortality (HR=0.93 per SD, 95% CI 0.88-0.97), but the relationship reversed during the pandemic: extroverted individuals appeared to suffer higher mortality than their introverted counterparts, although the effect was not significant (HR=1.20 per SD, 95% CI 0.93-1.54).

Extroversion was associated with greater pandemic-related excess mortality (HR=1.20/0.93=1.29 per SD, 95% CI 1.00-1.67). Compared with someone who scored at the mean level of extroversion, mortality rates prior to the pandemic were 10% lower for a person who was very extroverted (i.e., top 12% of the sample at Wave 1), while they were 12% higher for someone who was very introverted (i.e., 11^th^ percentile). In contrast, mortality rates during the pandemic appeared to be *highe*r for very extroverted individuals (HR=1.15, 95% CI 0.77-1.71) and l*ower* for those who were very introverted (HR=0.70, 95% CI 0.43-1.14) although the difference was not significant because of limited statistical power. In sum, the slight mortality advantage enjoyed by extroverts prior to the pandemic disappeared during the first 10 months of the COVID-19 pandemic. It remains to be seen whether that pattern continued into 2021-22. We suspect that the mortality benefit of introversion during the pandemic is largely a result of reduced exposure to the risk of infection, but it may also derive in part from the ability of introverts to adapt more easily to reduced social interaction without engaging in self-destructive behavior (e.g., drug and alcohol abuse). Introverts have been training for a pandemic their whole lives.

## INTRODUCTION

Is being an extrovert good for your survival? We think the answer depends on when you ask the question. Prior to the pandemic, some evidence suggests that the answer is probably a weak yes (Chapman et al., 2020; Graham et al., 2017; Wilson et al., 2005). Yet, the benefit might be offset to some degree if extroversion is also associated with harmful behaviors. One study reported higher mortality among extroverts, which they attributed primarily to higher levels of smoking (Ploubidis and Grundy, 2009). Two other studies reported no significant relationship between extroversion and mortality (Shipley et al., 2007; Weiss and Costa, 2005).

According to the health-behavior model of personality, the main mechanism through which personality affects mortality is by influencing one’s propensity to adopt health-promoting behaviors and avoid behaviors that are harmful (Friedman, 2000; Smith, 2006). For example, extroversion could benefit health by enhancing social relationships (Berkman et al., 2000; Roberts et al., 2007), which has been linked to lower mortality (Holt-Lunstad et al., 2010).

Yet, extroversion could become a detriment during an airborne pandemic that thrives on human contact. Social life changed abruptly when COVID-19 was declared a global pandemic on March 11, 2020. In the interest of reducing contagion, social interaction was severely curtailed when offices, non-essential businesses, and schools were closed, while large social gatherings such as cultural and sporting events were canceled. Even a trip to the grocery store seemed like risky endeavor to many people. We hypothesize that introverts were better able to adapt to reduced social interaction. We suspect they were more willing to limit social activities and avoid large gatherings of people, thereby lowering risk of exposure to SARS-CoV-2. Apart from exposure-to-risk, introverts may have also been better-equipped to cope with reduced social interaction while still maintaining healthy behaviors (e.g., physical activity) without resorting to risk-taking behavior such as substance abuse. If so, introverts would have experienced fewer adverse indirect effects of the pandemic than extroverts.

To our knowledge, no one has evaluated the effect of extroversion on excess mortality during the pandemic, although there has been considerable attention paid to subjective feelings of loneliness, anxiety, and depression. Most of those studies suggest that extroversion was associated with bigger increases in loneliness (Entringer and Gosling, 2021; Folk et al., 2020) and greater deterioration in mental health during the pandemic (Proto and Zhang, 2021; Rettew et al., 2021). Similarly, we expect that extroverts experienced more excess mortality during the pandemic than introverts.

In this paper, we use data for midlife Americans surveyed in 1995-96 with mortality follow-up through December 31, 2020 to investigate whether the relationship between extroversion and mortality changed during the COVID-19 pandemic. We hypothesize that there will be an inverse association between extroversion and mortality during the prepandemic period (1995-Feb 2020), but the relationship will be reversed during the pandemic (March-Dec 2020). That is, we anticipate that excess mortality during the pandemic will be greater for extroverts than for introverts.

## METHODS

### Data

The data come from the Midlife in the United States (MIDUS) study, which targeted non-institutionalized, English-speaking adults aged 25–74^1^ in the contiguous United States (Brim et al., 2020). At baseline (fielded January 1995–September 1996), national random digit dialing with oversampling of older people and men was used to select the main sample (*N*=3,487) and a sample of twin pairs (*N*=1,914). The study also included a random sample of siblings of individuals in the main sample (*N*=950) and oversamples from five metropolitan areas in the U.S. (*N*=757). The response rate for the phone interview ranged from 60% for the twin subsample to 70% for the main sample. Among those who completed the phone interview (*N*=7,108), 6,325 (89%) also completed mail-in self-administered questionnaires (SAQs).

### Measures

Vital status was ascertained through searches of the National Death Index, survey fieldwork, and longitudinal sample maintenance (Ryff et al., 2022). We analyzed deaths through May 31, 2020 (see S1 in Supplementary Material for details). Among those who completed the SAQ at baseline, 1,767 (27.9%) died by May 31, 2020.

Personality was measured at Wave 1 using the standardized questionnaire for the “Big Five” taxonomy of personality (John, 1990). Each personality trait was based on the degree to which the respondent endorsed a set of four to seven descriptors, using response categories that included “not at all” (1), “a little” (2), “some” (3), “a lot” (4) (Lachman and Weaver, 1997). Extroversion was computed as the mean across five descriptors (Outgoing, Friendly, Lively, Active, Talkative; α=0.78). Conscientiousness was measured by 4 items (Organized, Responsible, Hardworking, Careless [reverse-coded]; α=0.56). Neuroticism included 4 items (Moody, Worrying, Nervous, Calm [reverse-coded]; α=0.75). Agreeableness was based on 5 items (Helpful, Warm, Caring, Softhearted, Sympathetic; α=0.81). Openness comprised 7 items (Creative, Imaginative, Intelligent, Curious, Broad-minded, Sophisticated, Adventurous; α=0.78).

We included potential confounders that may affect personality and are known to be associated with mortality. Demographic confounders comprised sex, age, and race/ethnicity. Respondents were asked, “*What race do you consider yourself to be?*” We retained the first two response categories (i.e., Black and/or African American; White), but combined the remaining categories (i.e., Asian or Pacific Islander; multiracial; Native American or Aleutian Islander/Eskimo; other) into a group labeled “other race.” Ethnicity is based on reported countries of origin (“*Other than being American, what are your main ethnic origins? That is, what countries or continents are your ancestors from?*”). We classified respondents as Latino/a if they reported a country of origin in Mexico, Central America, Cuba, Dominican Republic, Puerto Rico, South America (including Brazil), or Spain. For respondents who were missing information regarding race/ethnicity from Wave 1 (2% of the sample), we used information from Wave 2: respondents were asked to identify the race with which they most closely identify (“*Which do you feel best describes your racial background? White, Black or African American, American Indian or Alaska Native, Asian, or Native Hawaiian or Pacific Islander?*”); Latino/Hispanic origin was also based on self-report (“*Are you of Spanish, or Hispanic or Latino descent, that is, Mexican, Mexican American, Chicano, Puerto Rican, Cuban or some other Spanish origin?*”). In sensitivity analyses, we also adjusted for self-assessed health status and physical limitations atbaseline. Self-assessed health status was measured on a five-point ordinal scale ranging from 1 (“poor”) to 5 (“excellent”). An index of physical limitations was based on self-reported limitations doing the following 8 tasks: lifting/carrying groceries; climbing several flights of stairs; bending/kneeling/stooping; walking more than a mile; walking several blocks; walking one block; vigorous activity (e.g., running, lifting heavy objects); and moderate activity (e.g., bowling, vacuuming). The response categories ranged from “not at all” (0) to “a lot” (3). Based on the recommendations of Long & Pavalko (2004), we constructed the index by summing the 8 items (potential range: 0–24), adding a constant (0.5), and taking the logarithm of the result, which allows for relative rather than absolute effects.

### Statistical Analysis

We used standard practices of multiple imputation to handle missing data (Rubin, 1996; Schafer, 1999); see S2 in Supplementary Material for more details. A Cox model was used to model age-specific mortality with a robust variance estimator to correct for family-level clustering. Age was treated as the time metric, but we also treated calendar year as a time-varying covariate to adjust for the period trend in mortality decline. Among the MIDUS cohort, mortality was likely to rise over time purely as a result of aging (i.e., the cohort aged 25 years between 1995 and 2020, which is why it is important to model age-specific mortality), but mortality also tends to decline over time among the population as a whole (i.e., the mortality rate for someone who was aged 50 in 1995 was likely to be much higher than the corresponding rate for their younger counterpart who reached age 50 in 2015). Between 1995 and 2019, life expectancy increased by 3.3 years, from 75.9 years in 1995 to 79.2 years in 2019 (University of California, Berkeley and Max Planck Institute for Demographic Research, 2022); correspondingly, the percentage of Americans dying before age 75 fell from 38% in 1995 to 30% in 2019. Thus, controlling for age and in the absence of a pandemic, we would have expected mortality to be substantially lower in 2020 than in 1995.

Model 1 adjusted for age (as the “clock”), sex, race/ethnicity, calendar year (i.e., to capture period mortality decline), a dichotomous indicator (P) for the pandemic period (which indicates whether mortality deviated from the prepandemic trend), extroversion (£), and an interaction between extroversion score and the pandemic indicator. Thus, we divided the survival history for each respondent into the intervals representing each calendar year from 1995 through 2019, the prepandemic portion of 2020 (i.e., January-February), and the final pandemic period (March-December 2020) in order to specify period as a time-varying covariate. We evaluated different specifications for the period trend (i.e., linear, quadratic, 5-year categories) based on mortality through 2019; the linear specification yielded the best fit (i.e., based on the Bayesian Information Criterion). Thus, we treated period as linear in the final models.

The pandemic indicator represents the extent to which mortality after March 2020 differed from the expected level of mortality in the absence of a pandemic after accounting for aging of cohort and period mortality decline. A hazard ratio (HR) greater than 1.0 implies excess mortality during the pandemic (i.e., mortality is higher than expected based on the prepandemic mortality linear trend), whereas a value less than 1.0 indicates that mortality was lower than expected. Excess mortality includes deaths resulting directly from COVID-19 (whether recorded as such or not) as well as potential increases in mortality from other causes indirectly affected by the pandemic.

To ease interpretation, we reparameterized the model to include two interaction effects for extroversion rather than a main and an interaction effect. The first interaction (*E* x (1 - *P*))) represents the effect of extroversion during the prepandemic period; it is the same as the main effect in a standard specification. For this interaction, we expected a hazard ratio less than 1.0, indicating that extroverts experienced lower mortality than introverts prior to the pandemic. The second interaction (*E* x *P*) represents the effect of extroversion during the pandemic period; the coefficient for this interaction equals the sum of the main effect and the interaction effect from the standard specification (i.e., the HR is the product of the HRs for the main and interaction effects). For this interaction, we expected a hazard ratio greater than 1.0, implying that extroverts experienced higher mortality than introverts during the pandemic.

Model 2 further adjusts for the main effect of conscientiousness, which is the personality trait previously reported to be the most strongly and consistently associated with mortality (Chapman et al., 2020; Graham et al., 2017; Iwasa et al., 2008; Jokela et al., 2013; Weiss and Costa, 2005). We might expect conscientious individuals to exhibit greater compliance with public health orders to socially distance, wear a mask in higher-risk settings, accept vaccination, and stay up-to-date with appropriate boosters. And, as expected, conscientiousness conferred a mortality advantage even before the pandemic. In auxiliary models, we tested an interaction between conscientiousness and the pandemic indicator, but found no evidence that the effect of conscientiousness differs significantly between the prepandemic and pandemic period. That is, conscientiousness continued to be associated with lower mortality throughout the period, but there was no indication that the mortality advantage increased during the pandemic. In Model 3, we added the other three personality traits (i.e., neuroticism, openness, and agreeableness). Finally, Model 4 further controls for self-assessed health status (treated as categorical) and the index of physical limitations.

The temporal ordering of personality and health status is unclear. In theory, one might expect personality to be a stable trait that remains relatively constant throughout life; however, research suggests that some facets of personality may evolve over the life course. For example, Roberts et al. (2006) found that one facet of extroversion, social dominance, increased with age during young adulthood but then levels out after age 35; another facet of extroversion, social vitality, showed little change with age. In this analysis, we used only the Wave 1 measures of personality traits. However, MIDUS measured the Big Five personality traits at all three survey waves, and there was some notable variation in the scores over time for a given individual.^2^ It is possible that the association between extroversion and mortality was confounded by health status (e.g., seriously ill respondents were less likely to be social active, which may have affected their reporting regarding extroversion). On the other hand, health status may also mediate the relationship between personality and mortality, in which case controlling for health status may under-estimate the total effect of personality.

### RESULTS

The cohort included Americans aged 20-75 at baseline (1995-96), 28% (*N*=1,767) of whom died by December 31, 2020 (Table 1). Survivors were aged 31-98 at the end of mortality follow-up. The mean score on extroversion was 3.2, with a distribution that was heavily weighted toward the extroverted end of the spectrum. Only 11% scored <2.4 (< 1.43 SD below the mean); 29% scored <2.4 (< 0.7 SD below the mean); 23% scored above 3.6 (> 0.7 SD above the mean); and 12% scored the maximum of 4 (1.42 SD above the mean). The mean score was highest for agreeableness (3.5) followed by conscientiousness (3.4) and lowest for neuroticism (2.2).

**Table 1.** Descriptive statistics for analysis variables, *N*=6,325^a^.

| Variable | Mean (SD) or Percent |
| --- | --- |
| Age at baseline<sup>b</sup> (20-75), mean (SD)<sup>b</sup> | 46.9 (12.9) |
| Male, % | 47.5 |
| Non-Latina/o White, % | 88.3 |
| Non-Latina/o Black, % | 5.2 |
| Non-Latina/o Other race, % | 2.7 |
| Latina/o, % | 3.8 |
| Extroversion (1-4), mean (SD) | 3.2 (0.6) |
| Conscientiousness (1-4), mean (SD) | 3.4 (0.4) |
| Neuroticism (1-4), mean (SD) | 2.2 (0.7) |
| Openness (1-4), mean (SD) | 3.0 (0.5) |
| Agreeableness (1-4), mean (SD) | 3.5 (0.5) |
| Self-assessed health status |  |
| Poor, % | 2.6 |
| Fair, % | 11.1 |
| Good, % | 33.6 |
| Very Good, % | 35.8 |
| Excellent, % | 16.9 |
| Index of physical limitations (-0.7 to 3.2), mean (SD) | 0.8 (1.3) |
| Died by 12/31/2020, % | 27.9 |

<sup>a</sup> With the exception of mortality, all variables are measured at baseline (1995-96).
<sup>b</sup> At the end of mortality follow-up (12/31/2020), survivors were aged 31-98.

Prior to the pandemic, extroversion was associated with somewhat lower mortality (HR=0.93 per SD, 95% CI 0.88-0.97; Table 2, Model 1). In contrast, the effect of extroversion reversed during the pandemic: extroverted individuals appeared to suffer higher mortality than their introverted counterparts, although the effect was not significant (HR=1.20 per SD, 95% CI 0.93-1.54). Given the relatively small number of deaths during March-December 2020 (*N*=79, 14 of which resulted from COVID-19), the confidence intervals are very wide for the pandemic period. Nonetheless, the results indicate that extroversion was associated with greater pandemic-related excess mortality (HR=1.20/0.93=1.29 per SD, 95% CI 1.00-1.67); that is, compared with introverts, extroverts suffered a bigger increase in mortality during the pandemic relative to their prepandemic mortality levels.

**Table 2.** Hazard Ratios (95% CIs) from Models Predicting Age-Specific Mortality, MIDUS, 1995-2020.

|  | (1) | (2) | (3) | (4) |
| --- | --- | --- | --- | --- |
| Male | 1.34***<br>(1.22 - 1.48) | 1.32***<br>(1.20 - 1.45) | 1.37***<br>(1.25 - 1.52) | 1.48***<br>(1.34 - 1.64) |
| Non-Latina/o White | 1.00 | 1.00 | 1.00 | 1.00 |
| Non-Latina/o Black | 1.39**<br>(1.11 - 1.74) | 1.38**<br>(1.10 - 1.73) | 1.39**<br>(1.11 - 1.75) | 1.18<br>(0.94 - 1.49) |
| Non-Latina/o Other race | 0.94<br>(0.67 - 1.33) | 0.87<br>(0.61 - 1.25) | 0.87<br>(0.61 - 1.24) | 0.71<br>(0.50 - 1.02) |
| Latina/o | 0.99<br>(0.73 - 1.35) | 0.99<br>(0.73 - 1.34) | 0.98<br>(0.72 - 1.33) | 0.90<br>(0.66 - 1.23) |
| Year – 1995<sup>b</sup> | 0.99***<br>(0.98 - 0.99) | 0.99***<br>(0.98 - 0.99) | 0.99***<br>(0.98 - 0.99) | 1.00<br>(0.99 - 1.01) |
| Pandemic (Mar-Dec 2020)<sup>c</sup> | 0.90<br>(0.70 - 1.16) | 0.90<br>(0.70 - 1.16) | 0.90<br>(0.70 - 1.16) | 0.89<br>(0.69 - 1.15) |
| Extroversion<sup>a</sup> during:<br>Prepandemic<sup>d</sup> | 0.93**<br>(0.88 - 0.97) | 0.97<br>(0.92 - 1.02) | 0.93*<br>(0.87 - 0.99) | 0.98<br>(0.92 - 1.04) |
| Pandemic<sup>e</sup> | 1.20<br>(0.93 - 1.54) | 1.24<br>(0.96 - 1.59) | 1.19<br>(0.92 - 1.53) | 1.25<br>(0.97 - 1.61) |
| Conscientiousness<sup>a</sup> |  | 0.86***<br>(0.82 - 0.91) | 0.86***<br>(0.82 - 0.91) | 0.92**<br>(0.87 - 0.97) |
| Neuroticism |  |  | 1.09**<br>(1.03 - 1.15) | 1.02<br>(0.97 - 1.08) |
| Openness<sup>a</sup> |  |  | 1.04<br>(0.98 - 1.10) | 1.07*<br>(1.01 - 1.14) |
| Agreeableness<sup>a</sup> |  |  | 1.07*<br>(1.01 - 1.13) | 1.01<br>(0.95 - 1.07) |
| Self-assessed health status |  |  |  |  |
| Poor |  |  |  | 1.00 |
| Fair |  |  |  | 0.60***<br>(0.47 - 0.77) |
| Good |  |  |  | 0.43***<br>(0.34 - 0.55) |
| Very Good |  |  |  | 0.33***<br>(0.26 - 0.43) |
| Excellent |  |  |  | 0.26***<br>(0.20 - 0.35) |
| Index of physical limitations<sup>a</sup> |  |  |  | 1.19***<br>(1.14 - 1.25) |

\* p<0.05, \*\* p<0.01, \*\*\* p<0.001
<sup>a</sup> Standardized; the hazard ratio represents the effect per SD.
<sup>b</sup> Represents the per year change in mortality relative to 1995.
<sup>c</sup> Represents the difference in mortality during the pandemic (Mar-Dec 2020) compared with the prepandemic period (1995 through Feb 2020) after adjusting for period mortality decline.
<sup>d</sup> Represents the effect of extroversion (per SD) on the mortality rate prior to the pandemic. We have reparameterized this model to include effects for extroversion during the prepandemic and during the pandemic periods rather than a main effect for extroversion and an interaction effect. This HR is the same as the main effect in the standard model specified with a main and interaction effect.
<sup>e</sup> Represents the effect of extroversion (per SD) on the mortality rate during the pandemic. This HR equals the product of the HRs for the main and interaction effects in a standard specification (i.e., the exponentiated sum of the coefficients). To obtain the HR for the interaction effect in the standard specification (i.e., the effect of extroversion on the level of excess mortality; that is, the degree to which the effect of extroversion differed between the prepandemic and pandemic periods), one can divide this hazard ratio by the corresponding hazard ratio prior to the pandemic.

After adjusting for conscientiousness (Model 2), the difference in the effect of extroversion between pandemic vs. prepandemic periods was only marginally significant (HR=1.24/0.97=1.28, 95% CI 0.99-1.65). After adjusting for the other three personality traits (Model 3) the effect of extroversion prior to the pandemic was somewhat stronger, whereas the effect during the pandemic was somewhat weaker. Nonetheless, the association between extroversion and excess mortality remained unchanged (HR=1.19/0.93=1.28, 95% CI 0.99-1.66).

To better demonstrate how the effect of extroversion changed during the pandemic, we computed the hazard ratios associated with selected levels of the extroversion score prior to and during the pandemic based on Model 3 (Figure 1). Compared with someone who scored at the mean level of extroversion, mortality rates prior to the pandemic were 10% lower for a person who was very extroverted (i.e., 1.42 SD above the mean, top 12% of the sample at Wave 1), while they were 12% higher for someone who was very introverted (i.e., 1.43 SD below the mean, 11^th^ percentile). However, the mortality advantage for extroverts disappeared during the COVID-19 pandemic. Although the differences are not significant (because of limited statistical power), the pattern of results suggests that, if anything, extroverts suffered *higher* mortality than introverts during the pandemic. Relative to those who scored at the mean level of extroversion, mortality rates during the pandemic appeared to be *highe*r for very extroverted individuals (HR=1.15, 95% CI 0.77-1.71) and *lower* for those who were very introverted (HR=0.70, 95% CI 0.43-1.14).

**Figure 1.**
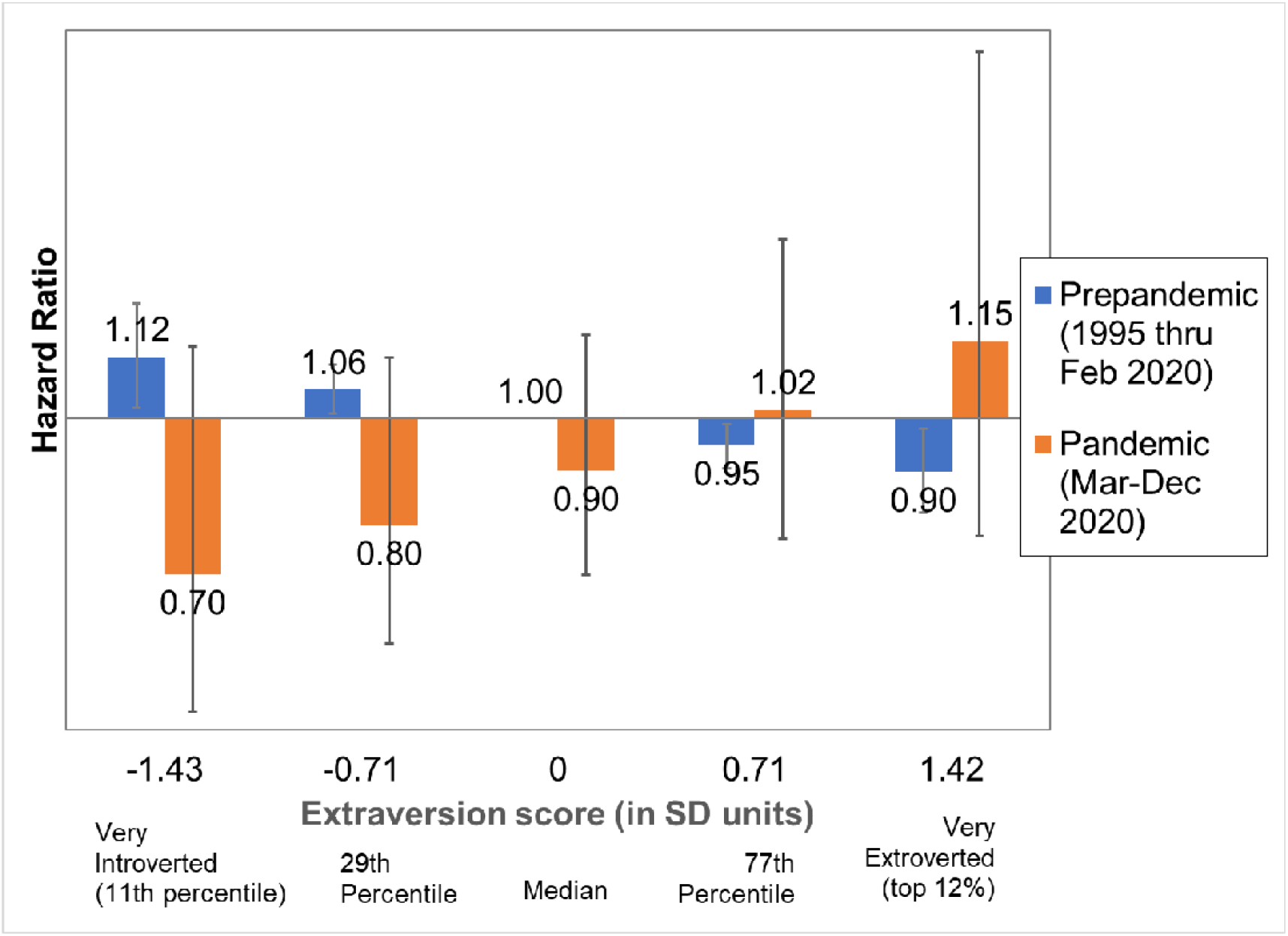
Hazard ratios for extroversion from model predicting age-specific mortality during prepandemic (1995 through Feb 2020) versus pandemic (Mar-Dec 2020) periods. Note. Based on a Cox model that uses age as the time metric and adjusts for sex, race/ethnicity, the linear period trend in mortality decline (prior to the pandemic), the main effects for all 5 personality traits, a dichotomous indicator for the pandemic period (Mar-Dec 2020), and an interaction between the extroversion score and the pandemic indicator, which tests whether the effect of extroversion differed between the prepandemic and pandemic periods.

When we translated the estimated mortality rates into survival ratios before versus during the pandemic (Figure 2), we found that the percentage expected to survive from age 25 to 85 fell 8 percentage points for someone who was very extroverted (from 57% to 48%), whereas it increased 15 percentage points (from 49% to 64%) for their very introverted counterparts. Thus, survival among extroverts during the pandemic was comparable with introverts prior to the pandemic, whereas introverts had even better survival during the pandemic than did extroverts prior to the pandemic.

**Figure 2.**
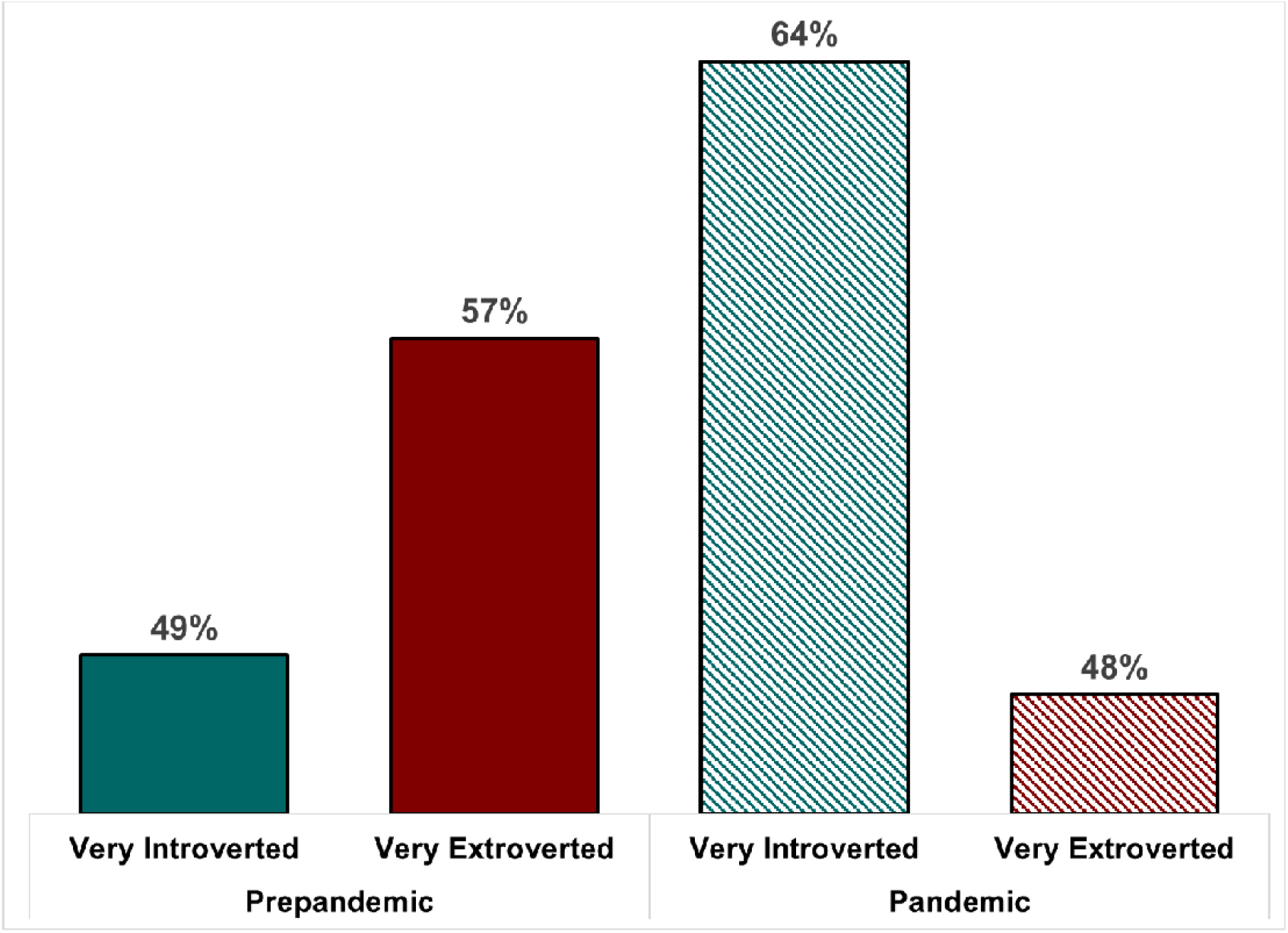
Fully-adjusted percentage surviving from age 25 to 85 by level of extraversion during prepandemic (1995 through Feb 2020) versus pandemic (Mar-Dec 2020) periods. Note: Estimates are based on Model 3 (Table 2) where the year is set to 2020, period is set to either prepandemic (Jan-Feb) or pandemic (Mar-Dec), and the extraversion score is fixed at the 11^th^ percentile (i.e., scored 2.4 out of 4, defined as very introverted) or the top 12% of the distribution (i.e., scored 4 out of 4, defined as very extroverted). All other covariates (i.e., sex, race/ethnicity, and the other four personality traits) are fixed at the mean for the sample.

### Sensitivity

When we further adjusted for self-assessed health status and physical limitations at baseline (Model 4), the hazard ratios for extroversion during the prepandemic period and for most of the other personality traits (except openness) were substantially attenuated. However, the hazard ratio for extroversion during the pandemic was, if anything, slightly stronger compared with Model 3 (albeit still not statistically significant).

## DISCUSSION

Introverts have been training for a pandemic their whole lives. As hypothesized, our results suggest that extroverts suffered higher excess mortality than introverts during the pandemic, although our statistical power is limited given that mortality follow-up is available only through 12/31/2020. We cannot say yet whether that pattern continued into 2021-22. The answer will have to wait until further mortality follow-up data become available.

Modern society is culturally biased towards extroverts, but a culture that favors extroversion and individualism is not the best prescription for surviving a pandemic. The mortality benefit of introversion during the pandemic was likely a result of reduced exposure to the risk of infection. Introverts may have been more willing to limit social activities, practice social distancing, and avoid large social gatherings, which would have made them less prone to deaths resulting directly from COVID-19.

Some of the benefit may also derive from the ability of introverts to adapt more easily to reduced social interaction. If they were less likely than extroverts to succumb to depression, anxiety, and/or loneliness during the pandemic as previous studies suggest (Entringer and Gosling, 2021; Folk et al., 2020; Proto and Zhang, 2021; Rettew et al., 2021), it could have suppressed mortality from other causes indirectly affected by the pandemic. If psychological distress contributed to excess mortality, we would expect to find an increase in suicide—the ultimate “death of despair.” Yet, there is little evidence that suicide rates increased during the pandemic. In fact, contrary to many predictions, suicide mortality was significantly lower than expected throughout March-December 2020 (Glei, 2022), although there appears to have been a small increase among Americans aged 25-34 (Ehlman, 2022).

In contrast, other so-called “deaths of despair” increased dramatically during the pandemic. Between 2019 and 2020, alcohol-related deaths increased 25% (White et al., 2022), while drug overdoses grew 30%, and in particular, deaths involving synthetic opioids such as fentanyl rose 55% (National Institute on Drug Abuse, 2022). If extroverts were more likely than introverts to succumb to substance abuse—perhaps because of difficulty coping with the stressors imposed by the pandemic—it could have led to more excess mortality from external causes.

### Limitations

There are several limitations to this study, First, mortality during 2020 is almost certainly under-estimated. Deaths during 2020 were based on an early release file for the National Death Index (NDI), which according to the National Center for Health Statistics, accounted for only about 95% of all recorded US deaths in 2020 at the time of the NDI search (Ryff et al., 2022). Second, the MIDUS sampling frame excluded the institutionalized population, who suffered especially high mortality early during the early stages of the pandemic. Thus, mortality among the MIDUS cohort is likely to be lower than pandemic-related mortality for the population as a whole. Third, we have no information about the degree to which MIDUS participants complied with public health orders during the pandemic and whether it differed by personality. Nor do we have any information about self-destructive behaviors (e.g., alcohol and drug abuse) during the pandemic. Fourth, personality was measured in 1995-96, approximately 25 years prior to the pandemic. Finally, the MIDUS sample under-represents minorities, who suffered higher mortality during the pandemic.

### Future Analyses

It will be useful to replicate this analysis using the Health Retirement Survey (HRS), which samples Americans older than 50, once mortality data become available for 2020. HRS has a much larger sample than MIDUS and thus, will yield more statistical power for modeling excess mortality. HRS also has a more ethnically diverse sample. There were only 14 deaths from COVID-19 among our MIDUS cohort, but 21% (N=3) of those deaths occurred to non-Hispanic Blacks whereas Blacks represented only 5% of survivors and 2% of those who died from some other cause during March-December 2020.

This analysis could also be repeated with non-US datasets such as the British Household & Panel Survey, the German Socio-economic Panel Study, and Household, Income and Labour Dynamics in Australia Survey, all of which were used in an earlier meta-analysis of the relationship between personality and mortality (Jokela et al., 2013). Prior to the pandemic, Jokela et al. (2013) found considerable variation across datasets in the association between extroversion and mortality: the inverse association was strongest in HRS followed closely by the Australian survey, but weaker and not significant in the UK, Germany, and the other two US survey (MIDUS and the Wisconsin Longitudinal Study, both of which have much smaller samples). It would be interesting to explore whether the relationship changed during the pandemic in other countries where the distribution of extraversion may be very different and where the pandemic response was less politicized. One study found that the US scored slightly higher on extroversion than Australia, but notably higher the UK and Germany (Kajonius and Giolla, 2017).

## Conclusion

The answer to the question posed at the start (“Is being an extrovert good for your survival?”) is: maybe…at least under normal conditions, but not in the midst of an airborne pandemic in which social contact could be deadly. Our results suggest that the slight mortality advantage enjoyed by extroverts under normal circumstances disappeared during the first 10 months of the COVID-19 pandemic. Even as we crossed the milestone of one million official COVID-19 deaths, most Americans were discarding their masks and returning to an unrestricted social life. We may want to remember some of the healthier practices learned during the pandemic because COVID is not over.

## Data Availability

All data produced are available online at https://doi.org/10.3886/ICPSR02760.v19 and https://midus-study.github.io/public-documentation/Mortality/Core/MIDUS_Core_MortalityCauseData_N2124_20220310.sav. The Stata do-files used for the analysis are available upon reasonable request to the authors.

https://midus-study.github.io/public-documentation/Mortality/Core/MIDUS_Core_MortalityCauseData_N2124_20220310.sav

## ACKNOWLEDGMENTS

This study was funded by the National Institute on Aging (P01 AG020166, U19AG051426) and the Graduate School of Arts and Sciences, Georgetown University. We are grateful to Kristen Harknett, Samuel H. Preston, Jon Mormino, and Jennifer Dowd for suggestions on an earlier draft of this manuscript.

## Supplementary Material

### S1. MORTALITY FOLLOW-UP

Vital status was based on: 1) searches of the National Death Index (NDI); 2) Wave 3 tracing (conducted between May 2013 and 2017), and 3) longitudinal sample maintenance.(Ryff et al., 2020) The most recent mortality file for MIDUS includes deaths that occurred as late as December 2021, but the most recent NDI search covered the period through December 31, 2020; thus, mortality after 2020 is likely to be incomplete. The NDI search was based on final death data for 1995-2019 and an early release data file for 2020.

### S2. MULTIPLE IMPUTATION

Among the 6,325 respondents included in the analysis, the predictors with the highest percentage of missing data were race/ethnicity (2.9%) and personality measures (0.9-1.0%). We used the “ice” command to perform multiple imputation. For the multiple imputation process, we used information for all the analysis variables as well measures of childhood socioeconomic status (SES), marital status, employment status, adult SES, smoking history, alcohol abuse, drug abuse, and physical limitations.

For continuous variables, departures from normality may result in implausible imputations when using the default draw method. Household income and wealth had a skewed distribution. Prior to imputation, we applied an inverse hyperbolic sine (IHS) transformation to those variables (Killewald et al., 2017). To ensure that imputed values were within the range of observed values, we used prediction matching for those variables and several others for which imputation generated out of range values (i.e., age at baseline, personality measures, education, occupational socioeconomic index score, and physical limitations).

For imputation of ordinal variables (e.g., perceived financial situation in childhood, self-assessed health status), we used an ordered logit model for imputation. We performed five imputations and then used the “mim” prefix command to re-estimate the model for each imputation and combine the five sets of estimates using Rubin’s rules (Royston et al., 2009).

## Footnotes

1 Although the sampling frame targeted adults aged 25-74, the final sample included a few respondents aged 20-24 (*N*=15) or aged 75 (*N*=4) at the baseline phone interview.

2 Among those with a valid extroversion score at all three waves (N=2490), 19% exhibited a decline of more than one SD between Wave 1 and 3 (i.e., >0.55 of a point on the scale ranging from 1-4), while 8% showed an increase of more than one SD. There is no evidence that the changes over time in extroversion are significantly associated with age at baseline, sex, or race/ethnicity, but there was a strong inverse association between the level of extroversion at Wave 1 and the change in extroversion between Waves 1 and 3 (i.e., suggesting that a large share of the variation may simply be measurement error). Also, those with better self-assessed health (SAH) status at baseline were less likely to exhibit substantial changes over time in the extroversion score, whereas those who were currently smoking at Wave 1 were more likely to exhibit a big decrease in extroversion by Wave 3. Individuals with more physical limitations were also more likely to have at least a small decrease in extroversion. Finally, those who exhibited a big decline in extroversion (i.e., more than one SD) between Waves 1 and 3 were more likely to die after Wave 3 than those with relatively stable values of extroversion net of sex, age, race/ethnicity, and SAH at baseline. Thus, we suspect that individuals who exhibited large deviations over time in extroversion may have had other underlying health issues that influenced the manifestation of their personality.

